# A Protocol for Identifying Priorities for Women+ Health in the Maritime Provinces using a Priority Setting Partnership Approach

**DOI:** 10.64898/2026.04.02.26350048

**Authors:** Justine Dol, Christine Pritchett, LeeAnn Larocque, James Bentley, Melissa Brooks, Annette Elliott Rose, Natalie O. Rosen, Emma Davies, Madhuri Yeluri, Meghan Gosse

## Abstract

**Background/Objectives:** Women+ (e.g., women and individuals assigned female at birth) experience disproportionate health risks and persistent gaps in access to care, despite regionally coordinated health systems. Women+ health research remains significantly underfunded and understudied, contributing to inequities in diagnosis, treatment, and outcomes. This study aims to collaboratively identify and prioritize the most pressing unanswered research questions related to women+ health in the Maritime provinces of Canada.

**Methods:** This study will use a modified Priority Setting Partnership (PSP) methodology based on the James Lind Alliance framework. A mixed-methods participatory approach will be used, including bilingual online surveys (French, English) and a one-day consensus workshop. Participants will include women+, healthcare professionals, researchers, policymakers, and the public residing in the Maritime provinces (Nova Scotia, New Brunswick, and Prince Edward Island). An initial survey will collect research uncertainties through open-ended questions. A second interim survey will rank verified uncertainties, followed by a facilitated workshop to achieve consensus on the Top 10 research priorities. Qualitative data will be analyzed using content analysis, and descriptive statistics will summarize participant demographics.

**Anticipated Results:** This project is expected to generate a collaboratively developed, evidence-informed Top 10 list of research priorities for women+ health in the Maritimes. The process will also identify thematic gaps in existing research and assess feasibility considerations to inform future study design and implementation.

**Conclusions:** By centering women+ voices and engaging diverse interest holders, this study will establish a shared regional research agenda to guide future research, funding, and policy initiatives for women+ health research.

## 1. Introduction

The Maritime provinces of Nova Scotia, New Brunswick, and Prince Edward Island comprise 5.1% of the Canadian population, of which 50.7% identify as women+ (1). Despite the relatively low population density, women+ in the Maritimes face disproportionate health risks compared to other Canadians, including the highest incidence of breast and lung cancers (2,3). Across Canada and the Atlantic provinces, the leading causes of death for women+ between 2019 and 2023 were cancer, cardiovascular disease, and cerebrovascular disease (4,5). Addressing these disparities requires a clear understanding of regional women+ health priorities.

In this study, we use the term “women+” to reflect a gender-inclusive and additive approach to women’s health research. This framing acknowledges that not all individuals who experience health conditions traditionally associated with women identify as women, while also maintaining the visibility of women as a population in health research. Our approach aligns with recommendations by Brotto and Galea, which emphasize balancing inclusivity with the importance of not reducing individuals to biological characteristics alone (6). The term women+ encompasses cisgender women as well as transgender men, non-binary, Two-Spirit, and other gender-diverse individuals who may experience health concerns traditionally associated with women’s health or linked to sex-based biological characteristics across the lifespan. This inclusive framing recognizes that health conditions affect women+ uniquely throughout the lifespan.

Despite the critical importance, women+ health remains understudied and underfunded. In Canada, only 7% of national health research funding is allocated to women+ health (7). Moreover, even when women+ are included in research, sex and gender are not consistently reported, contributing to ongoing gaps in knowledge and inequities in care (8). For example, funding for research on non-reproductive organs (e.g., brain, heart) is six to seven times higher than for reproductive organs (e.g., breast, uterus)(9). These gaps have real-world consequences for women+, including increased risks of misdiagnosis, ineffective treatments, adverse drug reactions, and poorer health outcomes (10).

Healthcare delivery in the Maritime provinces involves both local provincial systems, including Nova Scotia Health, Horizon/Vitalité (New Brunswick), and Health PEI as well as IWK Health, which provides specialized and tertiary care to women and children across the Maritimes. Despite this infrastructure, access to primary care remains a persistent challenge. Recent data indicate that approximately 65,000 Nova Scotians and over 33,000 Prince Edward Island residents are on provincial registries for a primary care provider (11,12), while in New Brunswick, about 27.5% of residents report not having a regular primary care provider (13), highlighting persistent access gaps across the region. Additionally, national estimates suggest that approximately 14.4% of Maritime residents report lacking a regular healthcare provider, and 11.5% report unmet health needs (14). Population-level data indicate that women report higher rates of unmet healthcare needs than men (15), and emerging evidence suggests that gender-diverse individuals may face additional barriers in accessing appropriate care (16). These gaps underscore the need for research that identifies the specific needs of women+ in this region.

In 2025, the IWK Foundation, which supports IWK Health, a tertiary women’s and children’s health center in Atlantic Canada, conducted a region-wide consultation survey gathering over 27,000 responses from women+ across the Maritimes (17). The consultation survey identified several commonly reported concerns, including systemic barriers to accessing primary and specialist care, long wait times, gaps in reproductive and menopause-related services, and challenges navigating the healthcare system (17). While the survey provided valuable insights into community experiences and perceived gaps in care, it was not designed to systematically identify and prioritize researchable questions across interest holder groups. Building on this work, the present study describes the protocol and methodological approach to identify and prioritize the top research questions for women+ health in the Maritimes. Using a systematic, evidence-informed priority setting approach, the process will reflect the perspectives of women+, caregivers of women+, researchers, healthcare professionals, policy makers, and members of the public, centering women+ voices to guide future research, funding, and policy decisions and to strengthen regional collaboration for women+ health.

## 2. Materials and Methods

### 2.1. Study Design

This study describes a protocol for identifying and prioritizing research questions related to women+ health in the Maritime provinces of Canada using a modified Priority Setting Partnership (PSP) approach informed by the James Lind Alliance (JLA) methodology (18,19). The approach is considered modified because some methodological adaptations were made to suit the regional context and study objectives, including the integration of an evidence and gap map review, inclusion of researchers in the priority setting, and a hybrid consensus workshop format. The PSP approach is designed to bring together individuals with lived experience, caregivers, healthcare professionals, policymakers, and other interest holders to collaboratively identify and prioritize unanswered research questions of shared importance. The process is outlined in Figure 1 below.

**Figure 1.**
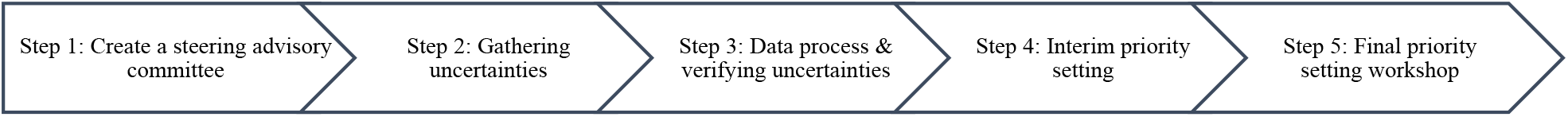
Priority Setting Process

### 2.2. Setting and Scope

The study will be conducted across the Canadian Maritime provinces of Nova Scotia, New Brunswick, and Prince Edward Island. Participants must reside in one of these provinces at the time of participation. While healthcare delivery models may differ across provincial systems, the PSP is designed to identify shared and regionally relevant research priorities in women+ health across the Maritimes.

The scope of the PSP focuses on women+ health across the adult life course and encompasses a broad range of physical, mental, emotional, social, and cultural health considerations. Areas of interest include, but are not limited to, reproductive and sexual health, chronic disease, mental health, perinatal and menopausal health, access to care, health system navigation, and experiences within healthcare settings. The PSP will focus on identifying researchable uncertainties rather than addressing individual clinical concerns or providing medical advice.

To promote awareness of this study, a website (www.maritimewomenshealth.com) provides information on the project and updates as the study progresses.

### 2.3. Data Collection Overview

The PSP consists of five sequential phases. Ethical approval from IWK Health Research Ethics Board has been obtained (REB#1032488).

#### Step 1: Create a steering advisory committee

This PSP will be overseen by a Steering Group comprising women+, healthcare professionals, researchers, and key interest holders in women+ health research in the Maritime Provinces. The steering committee is being led by first author (JD) and supported by a research coordinator (MG).

The Steering Group’s responsibilities include refining the PSP scope, providing input on the survey design and dissemination strategy, overseeing the categorization and organization of submitted questions, and supporting the final prioritization process. Members of the steering group were selected through a purposeful targeting of researchers, patient and public representatives, healthcare professionals, researchers, policy makers, and key organizations in women+ health research from across the Maritime Provinces.

#### Step 2: Gathering Uncertainties

To collect uncertainties, data will be gathered through a region-wide open-access online survey available to individuals residing in Nova Scotia, New Brunswick, and Prince Edward Island. All participants will review and provide informed consent electronically via REDCap (20) prior to participating in the study. Consistent with established Priority Setting Partnership methodology, no predefined sample size has been set; rather recruitment will focus on obtaining diverse perspectives across stakeholder groups and geographic regions (21). Rather, a pragmatic approach will be used, with survey recruitment focused on maximizing inclusivity across groups and provinces within available time and resource constraints. The survey will remain open for six weeks. Participants may choose to provide their email address if they wish to receive updates about the project or be involved in later stages of the PSP, including the interim ranking survey and/or final consensus workshop. Participant email addresses will not be linked to survey responses. Immediately following consent, participants will be directed to the online survey.

The survey has two components. First, the survey will collect demographic information to monitor representation across populations and interest holder groups. These data will be used descriptively to assess and describe the diversity of respondents and guide targeted recruitment efforts, rather than to make subgroup comparisons or draw conclusions about specific populations. Demographic items will include age, sex, gender identity, sexual orientation, education level, income, and province of residence. Participants will also be asked to self-identify their respondent group(s), which include a person with a lived experience as a woman+, a caregiver of a woman+, a friend or family member of a women+, a healthcare professional, a researcher, a policy maker, and/or a member of the public.

Second, the survey will solicit unanswered research questions related to women+ health in the Maritime provinces by asking participants to identify what questions about women+ health in the Maritimes they believe should be prioritized for future research. Participants will be informed that we are seeking questions related to all aspects of women+ health in the Maritimes, including physical, mental, emotional, social, and cultural factors. Participants may submit up to ten responses at a time and may complete the survey more than once. The survey will provide examples of women+ health related questions that focus on broader research priorities, including questions on physical, mental, emotional, social, or cultural health factors.

The survey will be administered online using REDCap (20) and will be available in both of Canada’s official languages (English and French). While this approach supports broad accessibility, it may limit participation from individuals who speak other languages, including some newcomer populations. Targeted recruitment through community organizations will be used to help mitigate this where possible.

Eligibility criteria and recruitment strategies are informed by equity, diversity, inclusion, and accessibility (EDIA) guidance and sex- and gender-based frameworks to promote diversity and inclusivity across age, gender identity, geography (urban and rural), province, cultural background, and lived experience, with the aim of ensuring broad representation of perspectives relevant to women+ health across the Maritime provinces (22,23). Recruitment material uses inclusive language to encourage participation from women+ and gender-diverse individuals, and targeted outreach will be conducted through community organizations and health networks serving diverse populations.

To support participation from historically underrepresented populations, outreach will also be conducted through organizations and community groups serving diverse populations across Nova Scotia, New Brunswick, and Prince Edward Island. Recruitment strategies will aim to engage these groups through targeted dissemination of the survey via healthcare networks, research institutions, community organizations, and advocacy groups across the Maritime provinces. This may include promotion through: (1) partner organizations and advisory networks (e.g., the Maritime SPOR SUPPORT Unit); (2) community-based organizations and health authorities across the Maritime provinces (e.g., Perinatal NB, IWK Health); (3) social media platforms (e.g., Meta); and (4) email outreach and newsletters. A curated list of regional organizations serving diverse and underrepresented populations (e.g., Indigenous, racialized, immigrant, low-income, and gender-diverse populations) has been developed to guide outreach and dissemination of the survey to promote participation from a range of perspectives. Efforts will also be made to reach women+ who do not have easy online access through dissemination via community and health organizations, including Menopause Society of Nova Scotia and Nova Scotia Native Women’s Association, for example. Recruitment materials will emphasize that the survey can be completed at any time to support participation from women+ who may be postpartum, caregiving for young children, or experiencing health-related fatigue. Dissemination through trusted community and health organizations will also allow individuals to access and complete the survey at a time that is convenient for them.

The study team acknowledges that, in the absence of formal partnerships with specific communities, the survey may not fully capture the priorities of all groups. Findings will be interpreted with attention given to principles of equity, inclusion, and data sovereignty. Survey responses will be monitored throughout the recruitment period to assess representation across respondent groups and inform additional outreach efforts if certain perspectives are underrepresented.

To help ensure data quality and reduce the likelihood of fraudulent or automated responses, the survey will incorporate security and verification measures (e.g., CAPTCHA and attention checks). Responses will also be reviewed during data cleaning for indicators of potential duplicate or invalid entries.

#### Step 3: Data process & verifying uncertainties

Following completion of the initial survey, all submitted questions will undergo a structured data cleaning, review, and refinement process conducted by the research team. Survey responses will be cleaned to ensure accuracy and consistency, and submitted questions will be reviewed to remove duplicate, overlapping, or out-of-scope submissions. Where appropriate, similar questions will be grouped thematically and refined into clear, researchable uncertainties using plain-language wording, while preserving the original intent of participant submissions. To maintain consistency and minimize bias, two members of the team will review all submissions independently, resolving differences through discussion or by involving the Steering Group when necessary. Submissions deemed out of scope will be documented with the reason for exclusion to maintain transparency and reproducibility.

In parallel with gathering uncertainties, the research team will undertake an evidence and gap review to identify published literature on women+ health in the Atlantic provinces (24). This process will provide direct evidence of existing research as it relates women+ health and will help inform the basis of existing evidence.

The refined list of questions will be reviewed against the evidence and gap map and existing literature to determine whether uncertainties have already been addressed by current research, both within the Maritimes and beyond. This process will focus on whether the specific uncertainty has been answered by existing research evidence, rather than whether a broader topic area has been studied. Questions identified as clearly addressed by existing research evidence will be removed, while those that remain partially or wholly unanswered will be retained. Determinations will be made through review by the research team and discussed with the Steering Group to ensure consistency and transparency. This process will result in a consolidated “long list” of verified research uncertainties related to women+ health in the Maritimes. Any research questions removed at this stage will be reported separately and justification of evidence will be noted for transparency.

#### Step 4: Interim priority setting

An interim prioritization survey will then be developed and disseminated using the same online platform (REDCap (20)) and recruitment strategies as the initial survey. Participants will be asked to review the long list of uncertainties and to rank or select the questions they believe are most important for future research. Uncertainties will be grouped by women+ health theme (e.g., cancer, perinatal, heart health) under which questions will be listed, and participants will be able to select the themes that they want to vote on. Participants will rank the questions within each theme in order of importance on a 5-point Likert scale from 1 (“not important”) to 5 (“very important”). At the end of this step, responses will be aggregated to produce a shortlist of 20–25 top research priorities per thematic area. These shortlisted uncertainties will then be compiled into a consolidated list to be considered during the final priority-setting workshop.

#### Step 5: Final priority setting workshop

The final phase will involve a facilitated one-day consensus workshop conducted using a hybrid format (in-person with a virtual participation option). Approximately 30 participants will be purposively selected to reflect diversity across interest holder roles (women+, caregivers, healthcare professionals, researchers, policymakers, and members of the public), as well as variation in gender identity, age, geography, and lived experience related to women+ health. We will also offer a virtual option for those who cannot travel to Halifax, Nova Scotia or if interest exceeds 30 individuals.

Workshop participants will be provided with the shortlisted research questions in advance of the meeting to allow for reflection and preparation. The workshop will be facilitated using participatory methods (e.g., journey mapping, dot-voting, empathy mapping) using a combination of small group work and full-team sessions to support diverse contributions for participants in-person and online (25). These approaches are commonly used in participatory research and priority setting processes to support inclusive dialogue and shared decision-making. These methods are designed to support equitable participation and ensure that all perspectives are heard, regardless of professional background or lived experience (26).

Through iterative discussion and ranking, participants will collaboratively review the consolidated shortlist of uncertainties and work toward reaching consensus on a final, ranked Top 10 list of research priorities for women+ health in the Maritime provinces. The outcome of this phase will be a set of community-endorsed research priorities that reflect shared decision-making and can be used to guide future research, funding, and policy initiatives in the region.

### 2.4. Data Management and Analysis

All survey data will be collected and managed using REDCap (Research Electronic Data Capture; Vanderbilt University (20)), a secure, web-based data capture platform approved by the host institution. Data will be stored on secure institutional servers, with access restricted to authorized members of the research team. Any identifying information (e.g., contact details provided for future engagement) will be stored separately from survey responses and will not be linked to individual submissions during analysis.

Qualitative responses from open-ended survey questions will be exported and analyzed using qualitative content analysis. Responses will be reviewed, coded, and grouped into thematic categories to identify distinct research uncertainties related to women+ health. This process will involve consolidation of overlapping or duplicate submissions while preserving the original intent and language of participants, consistent with PSP methodology. Qualitative data management and coding will be supported using NVivo software.

Quantitative demographic data will be analyzed using descriptive statistics to summarize participant characteristics and assess representation across interest holder groups and provinces. Where appropriate, responses from the interim prioritization survey will also be examined descriptively to explore patterns in priority rankings across interest holder groups and geographic regions. Statistical analyses will be conducted using IBM SPSS Statistics.

All analytic procedures, including criteria for determining in-scope and out-of-scope submissions and approaches to consolidating uncertainties, will be finalized prior to data analysis and documented to support transparency, reproducibility, and auditability. All findings will be reported according to the reporting guideline for priority setting of health research (REPRISE) guideline (27). Any protocol amendments related to analysis will be reported.

## 3. Expected Outcomes and Impact

This study is expected to produce a community driven, regionally relevant list of the top research priorities for women+ health in the Maritime provinces. By centering the perspectives of women+, healthcare professionals, researchers, policymakers, and the public, the project will generate a consensus-based, evidence-informed Top 10 list of research uncertainties that reflect the needs and lived experiences of the Maritime provinces.

The findings are anticipated to have many impacts, including, but not limited to, informing the development of future research agendas by highlighting high-priority questions that remain unanswered in women+ health and helping to ensure that studies address areas of greatest need and relevance. Further, this study will provide actionable insights for health system planners, policymakers, and healthcare organizations to align services and programs with community-identified priorities. This study also has the potential to strengthen partnerships and networks among women+, researchers, healthcare professionals, and decision makers, fostering ongoing collaboration and patient-oriented research. While this PSP is not setting out to identify unique differences in priorities across groups, the study will aim to ensure that the resulting priorities reflect a broad range of perspectives and lived experiences related to women+ health in the Maritime provinces. Collectively, the outputs of this study will provide a strategic foundation to guide future research, funding, and policy decisions, while demonstrating a replicable and inclusive approach to priority settings that could be adapted in other regions or health contexts.

Beyond generating the priority list for academic publication, the project will actively mobilize these findings through multiple pathways. Results will be shared with study participants and community partners through accessible lay summaries, infographics, and project updates distributed via the project website and partner organizations and networks. The findings will also be embedded in future grant applications and research programs led by academic and community partners and disseminated through targeted policy briefs and presentations to inform decision-makers within Nova Scotia Health, Horizon/Vitalité, Health PEI, and IWK Health. These activities aim to ensure that interest holders who contribute to the priority-setting process remain informed about how their input shaped the final priorities and how the results may influence future research and health system planning.

## 4. Conclusions

This protocol presents a systematic, transparent, and inclusive approach to identifying and prioritizing research questions in women+ health across the Maritime provinces of Canada. The resulting Top 10 research priorities are expected to: (1) Inform evidence-based research funding decisions; (2) Guide collaborative, patient-oriented research initiatives; (3) Support the development of equitable, regionally relevant women+ health research agendas; and (4) Strengthen networks and capacity for ongoing interest holder engagement in women+ health research. By centering diverse voices and systematically translating community-identified needs into actionable priorities, this study provides a replicable model for inclusive priority setting that can be adapted to other populations, regions, or areas of health research.

## Author Contributions

For research articles with several authors, a short paragraph specifying their individual contributions must be provided. The following statements should be used Conceptualization, J.D; methodology, J.D; investigation, all; data curation, all.; writing—original draft preparation, J.D., M.G.; writing—review and editing, all; project administration, J.D., M.G.; funding acquisition, J.D. All authors have read and agreed to the published version of the manuscript.

## Institutional Review Board Statement

The study will be conducted in accordance with the Declaration of Helsinki, and has been approved by the Institutional Review Board by IWK Health (protocol code #1032488 March 19, 2026) for studies involving humans.

## Informed Consent Statement

Informed consent will be obtained from all participants involved in the study.

## Data Availability Statement

This protocol paper does not report any data. Data sharing is not applicable to this article.

